# Microbiota development in early life mediates association between mode of delivery and vaccine responses

**DOI:** 10.1101/2021.09.01.21262898

**Authors:** E.M. de Koff, D. van Baarle, M.A. van Houten, M. Reyman, G.A.M. Berbers, F. de Heij, M.L.J.N. Chu, E.A.M. Sanders, D. Bogaert, S. Fuentes

## Abstract

The gut microbiota in early life, when critical immune maturation takes place, may influence the immunogenicity of childhood vaccinations. We assessed the association between mode of delivery, gut microbiota development in the first year of life, and mucosal antigen-specific immunoglobulin G (IgG) responses against pneumococcal and meningococcal conjugate vaccination at ages 12 and 18 months, respectively, in a prospective birth cohort of 120 infants. Birth by natural delivery was associated with higher IgG responses against both vaccines, which for the anti-pneumococcal IgG response could be explained by a gut microbial community composition with high abundances of *Bifidobacterium* and *Escherichia coli* in the first weeks of life. High *E. coli* abundance in the same period was also associated with higher anti-meningococcal IgG responses. Our results suggest that associations between mode of delivery and antibody responses to routine childhood vaccines are mediated by gut microbiota development.

## Introduction

Vaccination in early childhood is estimated to save millions of lives each year^1^. Vaccine-induced protection is mediated through a combination of innate, humoral and cellular immunity, and is often quantified by measuring antigen-specific antibody titers^2^. Large interindividual variation in antibody responses to vaccines administered in early life may limit vaccine effectiveness, leaving some fully vaccinated infants unprotected against serious infectious diseases^3^. Factors that influence vaccine responses include, among others, genetics, sex, perinatal characteristics like gestational age, birth weight, maternal antibodies, and feeding type, but also more general factors like geographical region (reviewed in ^4^). Recent research has shown that the gut microbiota, i.e. the sum of all microorganisms residing in the human intestinal tract, also plays a role in immune responses to vaccination^5–11^. This offers a potentially modifiable target to improve immunogenicity of childhood vaccines.

The gut microbiome is seeded at birth and rapidly develops over the first months of life under the influence of mode of delivery, breastfeeding, antibiotic administration and nutrition^12–15^. Timely exposure to specific microbes within the critical ‘window of opportunity’ in early infancy shapes the immune system^16–18^, including the B cell and immunoglobulin repertoire^19,20^. Microbial imprinting on the immune system may in turn explain part of the variation in vaccine responses. In support of this hypothesis, it has been shown that antibiotic-induced microbial perturbances in an infant mouse model led to impaired antigen-specific immunoglobulin G (IgG) responses against five common childhood vaccines^21^. Microbiota perturbance due to antibiotic exposure also resulted in impaired immune responses to seasonal influenza vaccination in healthy adults without pre-existing immunity, suggesting that primary responses are more sensitive to microbiota changes than recall responses^7^. In human infants, the composition of the microbial community pre-vaccination has been correlated with systemic immune responses to oral rotavirus vaccine, oral poliovirus vaccine, *Bacillus* Calmette-Guérin, hepatitis B, and tetanus vaccines^5,6,10,11,22,23^. However, the temporal relationship between 1. early-life exposures, 2. gut microbiota composition, and 3. subsequent childhood vaccine responses has not yet been studied.

Here, we demonstrate in a healthy birth cohort that mode of delivery-associated differences in early-life gut microbial colonization patterns are associated with antigen-specific IgG responses to pneumococcal and meningococcal conjugate vaccination in saliva. These findings are key for the design of intervention strategies that modulate the gut microbiota to enhance vaccine immunogenicity in infants.

## Methods

### Study design and participants

Fecal samples, saliva and questionnaires were collected from 120 healthy, full-term infants who participated in a prospective birth cohort study, where we previously reported a significant effect of mode of delivery on the gut microbiota in the first months of life^24^. Details on study design were previously published^24,25^. For the current analyses, we expanded our dataset with data and samples up to 18 months from 78 (65%) subjects, who participated in the follow-up study beyond the first year of life. Both parents provided written informed consent. Ethical approval was granted by the Dutch national ethics committee.

Study visits were conducted within 2 hours post-partum, 24-36 hours after birth, at 7 and 14 days and at 1, 2, 4, 6, 9, 12 months and, for those who participated in the follow-up study, 18 months of age. Saliva for antibody measurement was collected at the ages of 12 and 18 months. Fecal samples for gut microbiota profiling were collected by the parents prior to each visit, and were directly stored in the home freezer. Details on sample collection are in the appendix. Saliva and feces were transported on dry ice and stored at -80°C.

Extensive questionnaires including vaccination dates were collected. Infants were vaccinated according to the Dutch national immunization program (NIP). Ten-valent pneumococcal conjugate vaccine (PCV-10) was administered to infants born before September 2013 (52/120 participants) at the ages of 2, 3, 4, and 11 months, and to infants born from September 2013 (68/120 participants) at the ages of 2, 4, and 11 months due to changes in the NIP. Meningococcus type C (MenC) conjugate vaccination was administered at the age of 14 months.

### Laboratory procedures

Antigen-specific IgG against the capsular polysaccharides of pneumococcal vaccine serotypes 1, 4, 5, 6B, 7F, 9V, 14, 18C, 19F, and 23F was measured in saliva obtained at 12 months of age, and IgG against MenC polysaccharide in saliva obtained at 18 months of age. Antibodies were quantified using fluorescent bead-based multiplex immunoassays (MIA) as previously described^26–28^ (appendix). IgG concentrations below the lower limit of detection, which ranged from 0.08 ng/ml for pneumococcal serotype 4 to 0.37 ng/ml for pneumococcal serotype 14, and was 0.21 ng/ml for MenC, were set at half the lower limit of detection.

Laboratory processing and microbiota profiling of fecal samples was previously described^24^. In short, bacterial DNA was extracted using a combination of mechanical and chemical lysis methods, and was quantified by quantitative (q)PCR targeting the 16S rRNA gene^29^. The V4 hypervariable region of the 16S rRNA gene was amplified, and amplicon pools were sequenced on the Illumina MiSeq platform (Illumina, San Diego, CA) along with isolation and PCR blanks as negative controls. Sequences were processed in our previously described bioinformatics pipeline^30^. This resulted in an abundance-filtered data set containing only operational taxonomic units (OTU) that represented at least 0.1% of all reads in at least two samples (623/6690 OTUs). Taxonomic annotations of the 16S rRNA gene sequences were validated using shotgun metagenomics (subset of 20 samples), and species-specific qPCR (all samples)^24^.

### Statistical analyses

Data analysis was performed in R version 4.0.3. All statistical tests were two-tailed, and p-values below 0.050 or Benjamini-Hochberg adjusted p-values below 0.100 was considered statistically significant. All analyses were performed using log-transformed IgG concentrations, and were adjusted for time between vaccination and saliva collection using a second degree polynomial to account for the natural kinetics of the antibody response.

Concordance between IgG concentrations was evaluated using Spearman’s rank-order correlations. IgG geometric mean concentrations (GMCs) were compared using ANOVA with post-hoc Tukey tests. Associations between early-life host or microbiota characteristics and IgG concentrations were assessed using multivariable linear models.

Gut microbiota alpha diversity was assessed by the number of observed species, and the Shannon diversity index (*phyloseq*^31^). Stability of the microbial community composition over time was calculated as the Bray-Curtis (BC) similarity (1–BC dissimilarity) between consecutive samples within individuals. Dirichlet multinomial mixture models were used to group infants into community state types (CSTs) based on gut microbiota composition (*DirichletMultinomial*^32^) (appendix). Differences in the gut microbial community composition according to CST were evaluated using permutational analysis of variance (PERMANOVA) (*vegan*^33^). Smoothing-spline analysis of variance (*metagenomeSeq*-package^34,35^) was used to detect differences in individual OTU abundances (present in at least 10% of samples) over time between infants with responses above and below the median antigen-specific IgG concentration. This method detects differentially abundant OTUs, and identifies the time intervals in which significant differences exist.

## Results

We investigated associations between early-life exposures, gut microbiota development in the first year of life, and subsequent antibody responses against pneumococcal and meningococcal conjugate vaccination in 120 healthy, full-term infants. Basic, lifestyle and environmental characteristics were previously published^24^, and are briefly summarized in **Table 1**. Follow-up of the infants and sample inclusion for IgG measurement are shown in **Figure S1**. Serotype-specific anti-pneumococcal IgG concentrations in saliva were measured in 101/120 (84.2%) infants at the age of 12 months (median 28 days [IQR 21-33] after the PCV-10 booster dose). Anti-MenC IgG concentrations in saliva were measured in 66/78 (84.6%) infants at the age of 18 months (median 116 days [IQR 105-120] after MenC vaccination). Geometric mean concentrations (GMC) of anti-pneumococcal IgG ranged between 7.33 ng/ml (95% CI 5.75-9.33 ng/ml) for serotype 23F and 27.30 ng/ml (95% CI 22.14-33.67) for serotype 19F. The anti-MenC IgG GMC was 10.64 ng/ml (95% CI 8.64-13.11 ng/ml) (**Figure 1A**). IgG concentrations against the 10 pneumococcal vaccine serotypes strongly correlated with each other (Spearman’s rho 0.53-0.84, adjusted p<0.001 for all pairwise correlations), and not with anti-MenC IgG antibodies (Spearman’s rho 0.14-0.30, adjusted p>0.469 for all pairwise correlations) (**Figure 1B**). As serotype-specific anti-pneumococcal IgG concentrations were strongly correlated, we focused our analyses on serotype 6B, which shows relatively weak antigenic properties, and is commonly found during (severe) pneumococcal disease^36^. Significant findings were validated for the other serotypes.

**Table 1.**
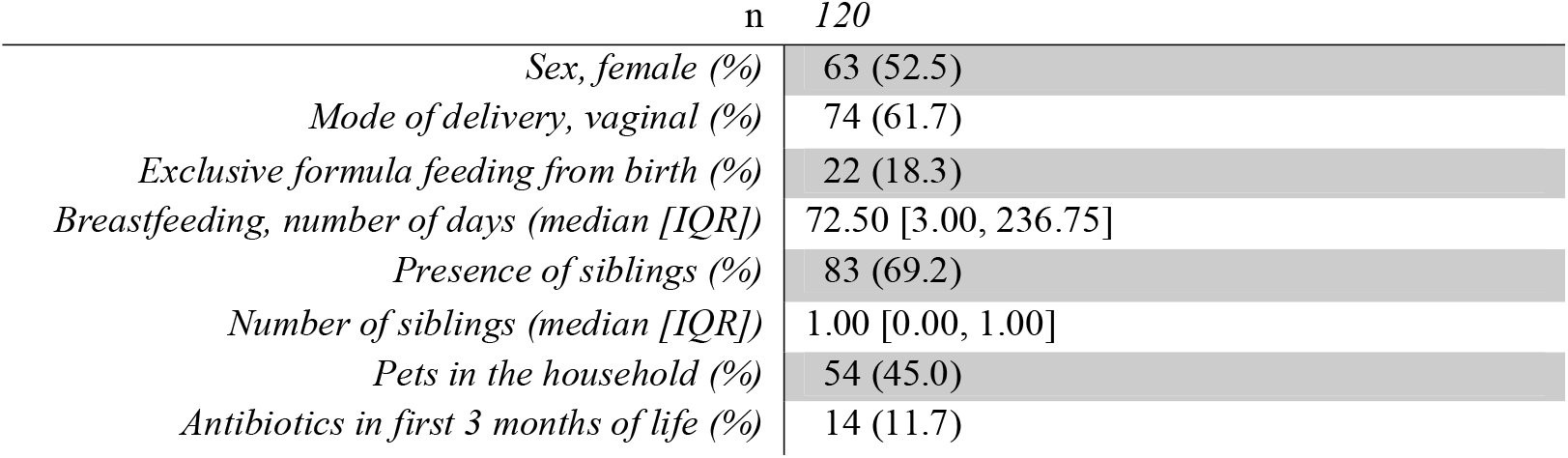
Cohort description.

**Figure 1.**
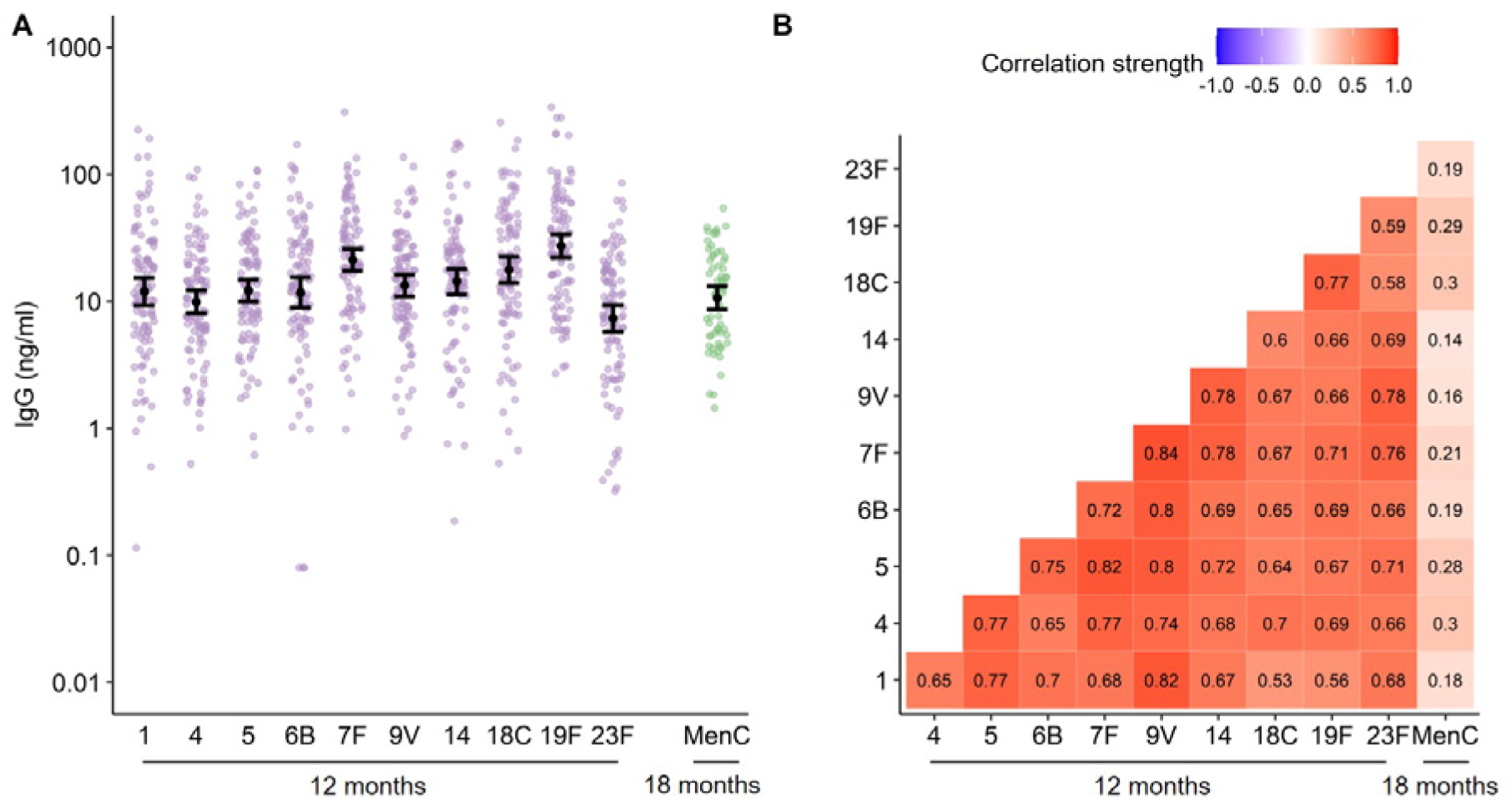
Anti-pneumococcal and anti-MenC IgG concentrations following vaccination. A) IgG concentrations against 10 pneumococcal vaccine serotypes (1, 4, 5, 6B, 7F, 9V, 14, 18C, 19F and 23F) and meningococcus type C (MenC) following vaccination. Black dots and error bars represent geometric mean concentrations with 95% confidence intervals. B) Correlation plot of IgG concentrations against the 10 pneumococcal vaccine serotypes and against MenC following vaccination. Numbers indicate the correlation strength, which was evaluated using Spearman’s correlation coefficients.

### Mode of delivery was associated with vaccine responses

We first investigated whether early-life host characteristics previously associated with differences in gut microbiome development and/or vaccine immunogenicity, were related to anti-Ps6B and anti-MenC IgG responses. Mode of delivery, feeding type, sex, antibiotics use in the first 3 months of life, and pets in the household were related to vaccine responses against one or more serotype, while having older siblings, and daycare attendance were not. These variables were included in multivariable linear models, including an interaction term between mode of delivery and feeding type due to the interdependency of these variables. Natural delivery (in contrast to caesarean (C-)section birth) was independently associated with higher anti-Ps6B IgG concentrations (β=0.51 [95% CI 0.043-0.97], p=0.033; **Figure 2A**). However, we also observed a negative interaction between natural delivery and exclusive formula feeding on anti-Ps6B responses (β=-1.32 [95% CI -2.43 - -0.21], p=0.021), suggesting that the positive effect of natural birth was diminished by subsequent formula feeding. Similar associations were found for IgG responses to most of the other pneumococcal vaccine serotypes (**Table S1**). Stratified analyses confirmed that, within the breastfed group, the anti-Ps6B IgG GMC of naturally born infants (n=51) was two-fold higher compared to C-section born infants (n=33; adjusted p=0.067); similarly, within the naturally born group, the anti-Ps6B IgG GMC of breastfed infants (n=51) was 3.5-fold higher compared to formula fed infants (n=7; adjusted p=0.083). Anti-Ps6B IgG concentrations did not differ between feeding types within the C-section born group (**Figure 2B**).

**Figure 2.**
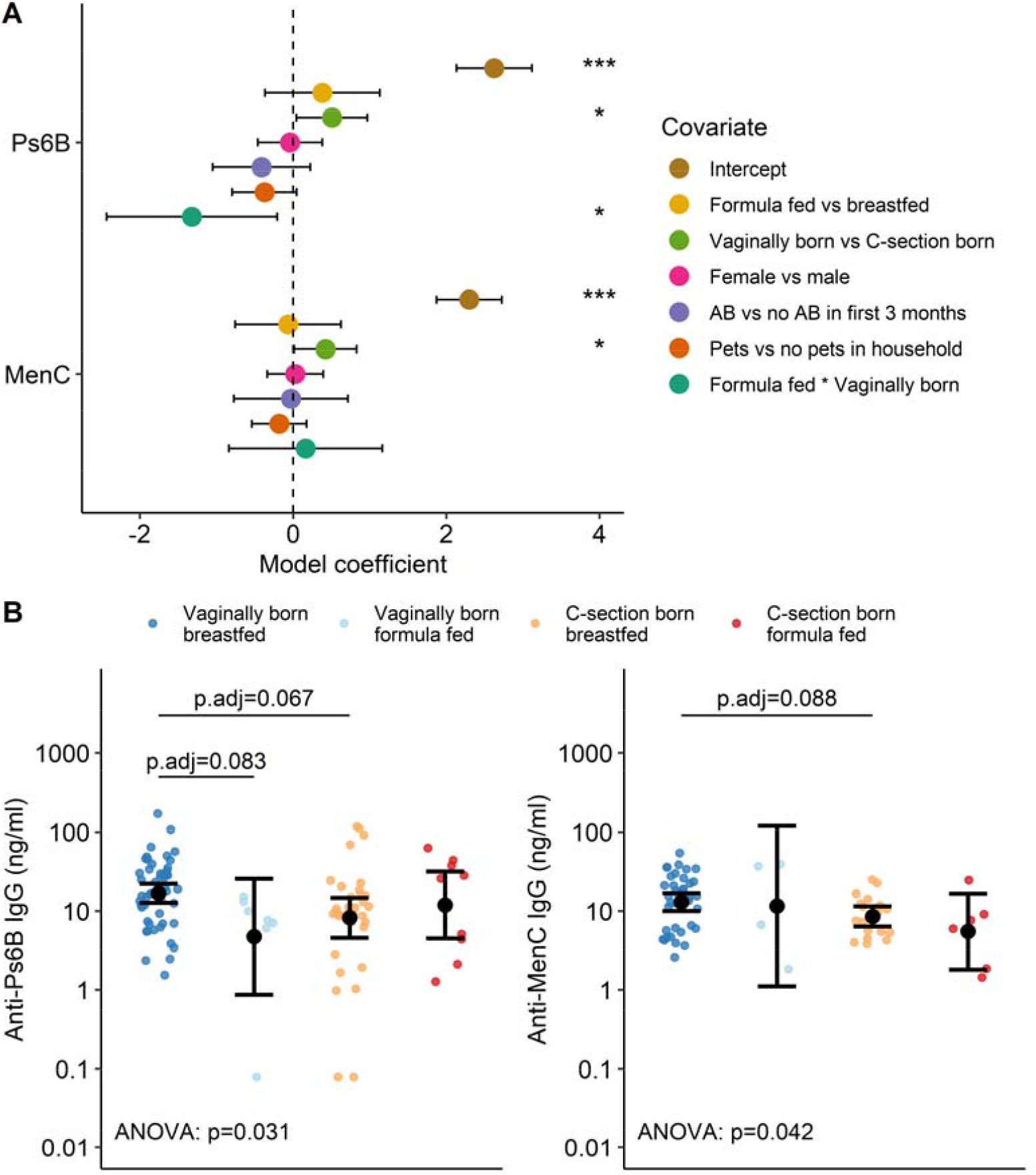
Associations between early-life exposures and anti-pneumococcal and anti-MenC IgG concentrations following vaccination. A) Colored dots and error bars represent coefficients with 95% CI of a multivariable linear model with log-transformed IgG concentrations as dependent variable. Significance is indicated by *: p<0.05; **: p<0.005; or ***: p<0.001. *Abbreviations:* C-section=caesarean section; AB=antibiotics. B) post-vaccination anti-pneumococcal serotype 6B (anti-Ps6B) IgG concentrations (left) and post-vaccination anti-meningococcus type C (anti-MenC) IgG concentrations (right) in infants stratified according to mode of delivery (vaginal birth vs. C-section) and feeding type (breastfeeding vs. exclusive formula feeding from birth). Black dots and error bars represent geometric mean concentrations with 95% CI. Significance was assessed using analysis of variance (ANOVA) on log-transformed IgG concentrations followed by a post-hoc Tukey test adjusting for multiple comparisons using the Benjamini-Hochberg procedure, also correcting for time between vaccination and IgG measurements.

Likewise, for MenC, natural delivery was also associated with higher IgG concentrations compared to C-section delivery (β=0.42 [95% CI 0.016-0.83], p=0.042), which was independent of feeding type (**Figure 2A**). In a stratified analysis of breastfed infants, naturally born infants (n=38) showed a 1.5-fold higher anti-MenC IgG GMC compared to C-section born infants (n=18; adjusted p=0.088; **Figure 2B**). Sex, antibiotic use, and having pets were not significantly associated with IgG responses against Ps6B or MenC.

### Gut microbial community composition at 1 week of age was associated with vaccine responses

We then studied whether gut microbiota development in the first year of life was associated with anti-Ps6B and anti-MenC IgG responses. Overall, 1052 out of 1177 fecal samples (89.4%) passed quality control for 16S rRNA gene-based sequencing, and were included in further analyses as previously described^24^.

No association was found between alpha diversity and anti-Ps6B or anti-MenC IgG concentrations at any time point, with the exception of an inverse correlation between the observed number of species at the age of two months and anti-Ps6B IgG concentrations (β=-0.029 [95% CI -0.049--0.0087], adjusted p=0.082). This association was not observed for the other pneumococcal vaccine serotypes (data not shown).

Gut microbiota stability between day one and week one, and between week one and week two correlated with higher anti-Ps6B IgG concentrations (day one-week one: β=1.66 [95% CI 0.44-2.88], adjusted p=0.074; week one-week two: β=1.22 [95% CI 0.22-2.22], adjusted p=0.077), which was not observed for any other time interval. Microbiota stability in the first two weeks of life was also significantly positively associated with IgG concentrations against all other pneumococcal vaccine serotypes (adjusted p≤0.083, **Table S2**). In contrast, no significant associations were found between microbiota stability and anti-MenC IgG concentrations.

The first two weeks of life, where gut microbiota stability was associated with anti-pneumococcal IgG concentrations, is compatible with the time frame when we previously found the largest difference in gut microbial composition between naturally born and C-section born infants in this cohort (at the age of one week)^24^. Therefore, we decided to focus on the microbial community composition in ‘week one’ samples, where we identified three distinct community state types (CSTs) (**Figure S2**). These CSTs differed considerably in community composition (PERMANOVA: R^2^=34.8%, p<0.001). Infants with CST1 (n=55) had a microbial community with low abundances of both *Bifidobacterium* and *Escherichia coli*, while infants with CST2 (n=48) had profiles with high *Bifidobacterium* abundances, and infants with CST3 (n=16) had high *E. coli* abundances (**Figure 3A**). Species-level microbial composition of the CSTs was largely confirmed by shotgun sequencing of 20 week one samples (**Figure S3**).

**Figure 3.**
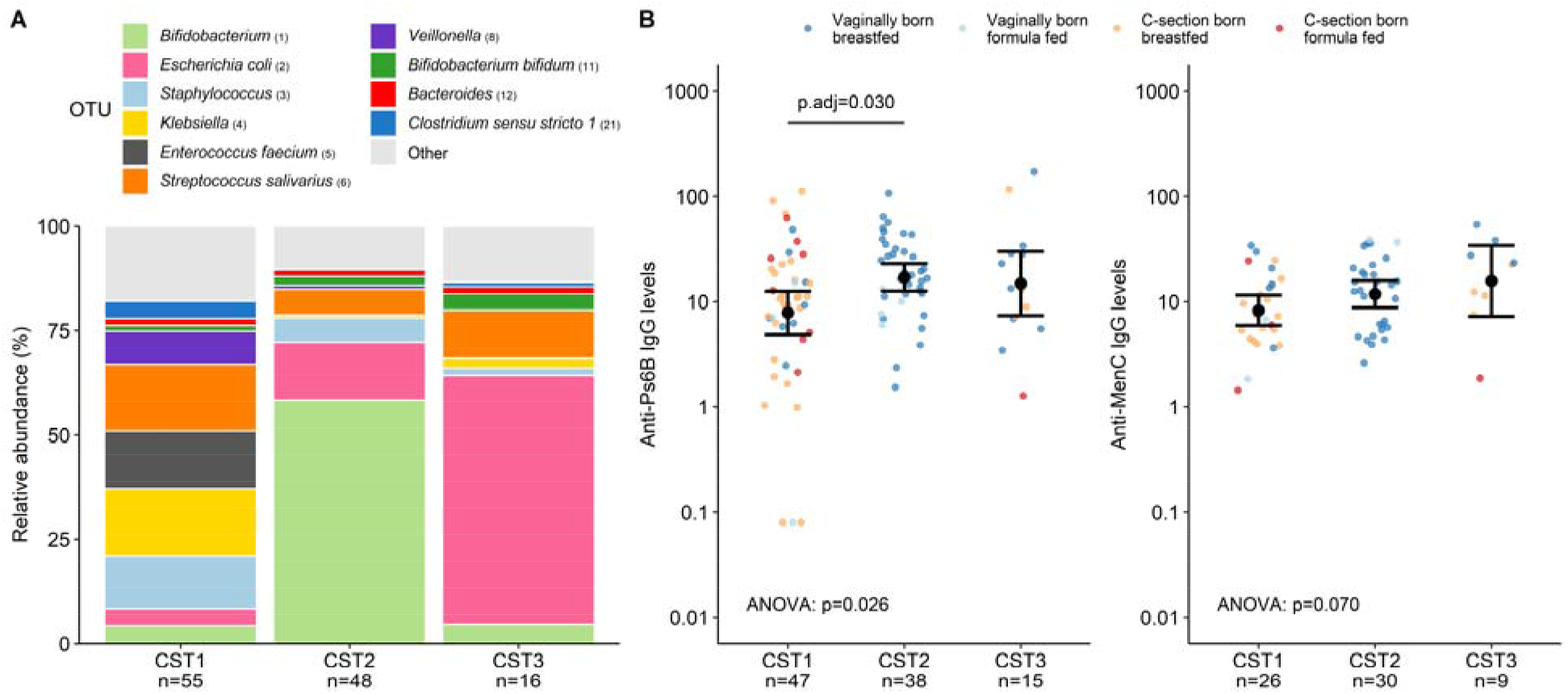
Gut microbial community state types at week 1 and anti-Ps6B and anti-MenC IgG concentrations. (A) Relative abundances of the top 10 OTUs per community state type (CST) defined at 1 week of age. (B) CSTs are plotted against anti-Ps6B IgG concentrations as representative pneumococcal serotype (left) and anti-MenC IgG concentrations (right). Dots are colored according to mode of delivery and feeding type from birth. Black dots and error bars represent geometric mean concentrations with 95% confidence intervals. Significance was assessed using ANOVA on log-transformed IgG concentrations followed by post-hoc Tukey tests adjusting for multiple comparisons using the Benjamini-Hochberg procedure, also correcting for time between vaccination and IgG measurements. *Abbreviations*: OTU = operational taxonomic unit; p.adj = adjusted p-value.

We then studied whether these CSTs were associated with anti-Ps6B and anti-MenC IgG concentrations following vaccination. Infants with CST1 had the lowest IgG concentrations against both Ps6B and MenC (anti-Ps6B IgG: GMC 7.84 ng/ml [95% CI 4.88-12.60]; anti-MenC IgG: GMC 8.28 ng/ml [95% CI 5.93-11.56]) (**Figure 3B**). Compared to infants with CST1, anti-Ps6B IgG concentrations were approximately two-fold higher in infants with CST2 (GMC 17.05 ng/ml [95% CI 12.64-23.00], adjusted p=0.030) as well as in infants with CST3 (GMC 14.85 ng/ml [95% CI 7.36-29.97], adjusted p=0.273), though only the comparison of anti-6B responses between CST1 and CST2 infants was significant. We observed similar associations between week one CSTs and IgG responses against most other pneumococcal vaccine serotypes (**Table S3)**. By contrast, anti-MenC IgG concentrations were not significantly different between CST groups, although infants with CST3 showed a nearly two-fold higher anti-MenC GMC (15.76 ng/ml [95% CI 7.25–34.26], adjusted p=0.148) than infants with CST1.

Mode of delivery was a strong driver of week one CSTs. All infants with CST2 were naturally born, which was significantly more than infants with CST1 (29.1%; Fisher’s exact test, adjusted p<0.001), or CST3 (62.5%, adjusted p<0.001). Natural birth was also overrepresented in infants with CST3 compared to CST1 (adjusted p=0.020). In contrast, feeding type (breastfeeding vs. exclusive formula feeding) was not significantly different between these CSTs. Interestingly, a post-hoc analysis revealed that the association between mode of delivery and anti-Ps6B IgG responses disappeared with the addition of week one CST as an independent variable, indicating that the positive effect of natural delivery on anti-Ps6B IgG depended fully on the CST. In contrast, natural delivery remained significantly associated with anti-MenC IgG responses, regardless of week one CST, suggesting an independent effect (**Table S4**).

To evaluate whether observed differences in early-life microbial community composition were sustained for a prolonged time, including time points closer to vaccination, temporal development of the gut microbiota according to week one CST was assessed using PERMANOVA. The microbial community composition of children according to their CST defined at week one converged over time, resulting in no differences between samples belonging to the CST groups from month six onward (**Figure 4A**). In pairwise comparisons, the observed differences in microbial community composition disappeared between infants with CST1 and CST3 by month one, between infants with CST2 and CST3 by month four, and between infants with CST1 and CST2 by month six. Similarly, relative abundances of *Bifidobacterium* and *E. coli* converged over time between CST groups (**Figure 4B**). At the age of 12 months, we identified two distinct CSTs, which were not significantly associated with anti-Ps6B or anti-MenC IgG responses, confirming that early-life microbiota were more strongly related to vaccine responses than the microbiota close to time of vaccination (**Figure S4**).

**Figure 4.**
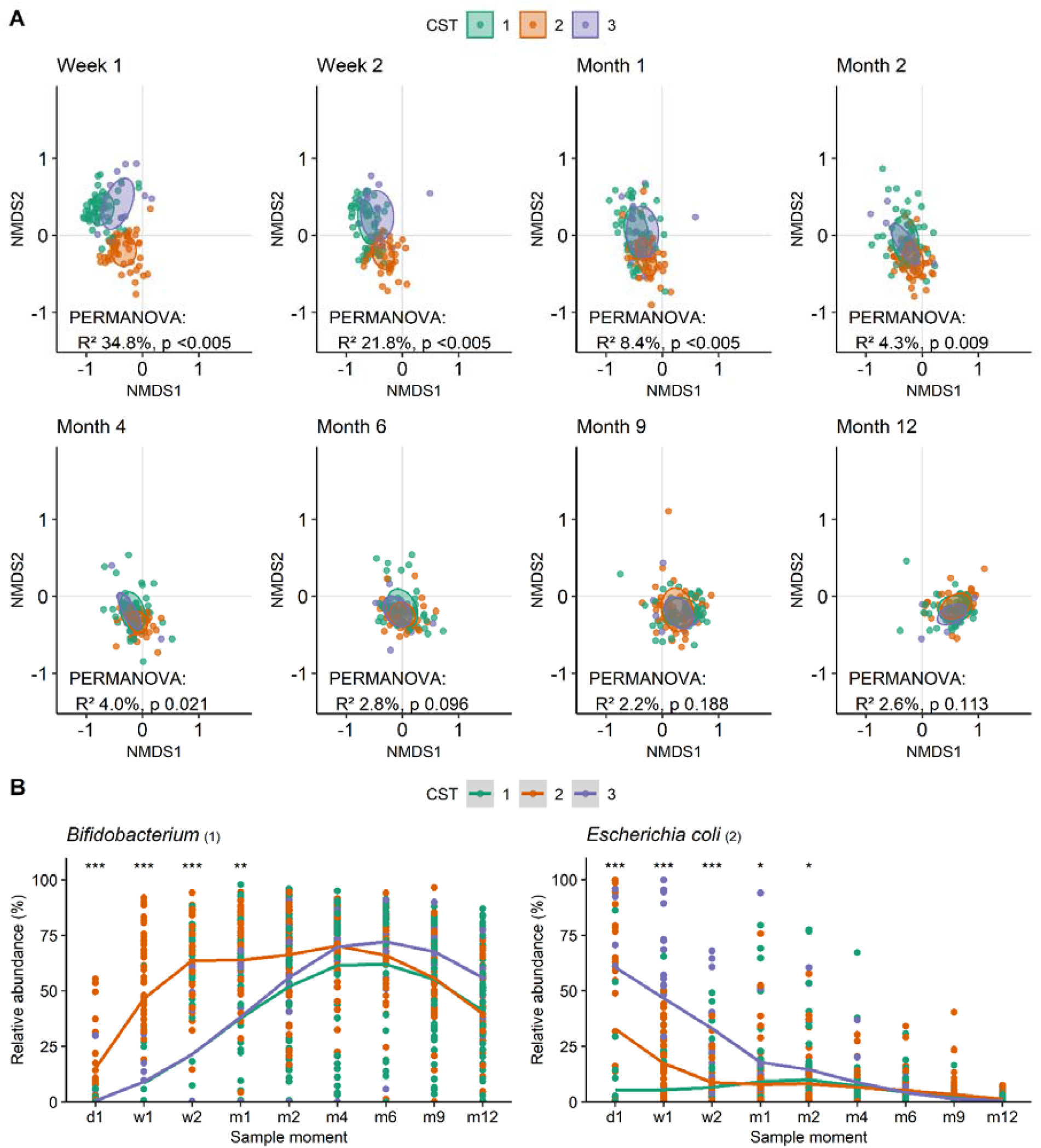
Temporal gut microbial composition development according to week 1 CST. (A) Non-metric multidimensional scaling plots based on Bray-Curtis dissimilarity, depicting the gut microbial composition per timepoint. Each dot represents the microbiota composition in a single participant’s sample. Infants are stratified according to week 1 community state type (CST). Ellipses represent the standard deviation of data points for each CST. Significance of differences according to week 1 CST was assessed using permutational analysis of variance (PERMANOVA). (B) Relative abundances of *Bifidobacterium (1)* (left) and *Escherichia coli (2)* (right) over time according to week 1 CST. Significance of differences according to week 1 CST was assessed using Kruskal Wallis tests and indicated by: *** for p<0.001, ** for p<0.005, and * for p<0.05.

### Early-life dynamics of individual OTUs are related to vaccine responses

Finally, we investigated differences in individual OTU succession patterns within the first two months between high and low vaccine responders (stratified along the median antigen-specific IgG response). Higher abundances of *E. coli* (days 0-41, adjusted p=0.013) and *Bifidobacterium* (days 0-5, adjusted p=0.027) were associated with high anti-Ps6B responses (confirmed for 7/9 other pneumococcal vaccine serotypes, **Table S5**). This was also observed for several *Bacteroides* OTUs, whereas *Clostridium, Prevotella* and *Streptococcus pyogenes* were associated with low responses (adjusted p<0.050).

Higher *E. coli* abundance (days 0-13, adjusted p=0.072) was also associated with high anti-MenC responses (**Table S6**). Because the MenC vaccination is administered at the age of 14 months, which is much later in life than the pneumococcal vaccinations, we extended the analysis to 12 months to allow for identification of associations with OTUs that colonize later in life. In high anti-MenC responders, we observed significantly higher abundances of multiple low abundant OTUs belonging to the Lachnospiraceae family, including *Fusicatenibacter saccharivorans* (days 101-381, adjusted p=0.080), *Pseudobutyrivibrio* (days 125-381, adjusted p=0.036) and several *Blautia* and *Roseburia* OTUs (**Table S7**).

## Discussion

We studied interactions between early-life exposures, gut microbial community development in the first year of life, and subsequent antibody responses in saliva against pneumococcal and meningococcal conjugate vaccination in a healthy birth cohort. A stable gut microbial community with high abundances of potentially beneficial bacteria in the first weeks of life, including *Bifidobacterium* and *E. coli*, was associated with high antibody responses to pneumococcal vaccination at 12 months of life. Furthermore, high *E. coli* abundance in early life was associated with high antibody responses to meningococcal vaccination at 18 months of life. Natural delivery was associated with high antibody responses to both vaccines, and, as we previously showed in this cohort^24^, with the early-life gut microbiota colonization patterns that we now associated with high antibody responses. Previous studies on associations between gut microbiota composition and serum antibody responses have focused on the microbiota near the time of vaccination^5,6,10,11,22^. However, our findings suggest that especially early-life gut microbiota development may set the stage for immune responses to childhood vaccinations.

Antibody responses to vaccination are elicited through activation of vaccine-specific B cells which will differentiate into immunoglobulin-secreting plasma cells and memory B cells. The gut microbiota have been implicated in the shaping of the systemic B cell and immunoglobulin repertoire in early life^19,20,37^. For instance, a deficient production of IgA and IgG1 in germ free mice can be restored by microbial exposure^38^. In infants, a culture-based study showed that the presence of *E. coli* and bifidobacteria in the gut in the first weeks of life was related to higher numbers of circulating CD27^+^ memory B cells at four and 18 months of life^39^. This suggests that bacterial colonization patterns in early infancy drive B cell maturation, and have a lasting effect on immunity^39^. In line with this observation, we found associations between gut microbiota community state types (CSTs) characterized by high abundances of *E. coli* and/or *Bifidobacterium* in 1-week-old infants and higher antibody responses to vaccination months later in childhood.

Previous studies have provided evidence for a positive effect of *E. coli* and *Bifidobacterium* on the immune response to vaccination. For instance, higher abundances of Gram-negatives including *E. coli* were associated with an adequate immune response against oral rotavirus vaccines^5^. Another study showed that treatment with the probiotic *E. coli* Nissle in a pig model enhanced the immune response to human rotavirus infection^40^, providing a causal link. A potential mechanism whereby *E. coli* may influence vaccine responses was pinpointed by a study demonstrating that impaired antibody responses to seasonal influenza vaccination in germ-free or antibiotic-treated mice were restored through TLR5-signaling by flagellated, but not unflagellated, *E. coli*^8^, suggesting strain- and antigen-specific immune enhancement. Furthermore, early-life absence of *Bifidobacterium* has been associated with reduced systemic immune responses to *Bacillus* Calmette-Guérin, polio virus, tetanus and hepatitis B vaccination^11,22^, which we also found for pneumococcal conjugate vaccination. Lack of early bifidobacterial colonization has been linked to immune dysregulation at the age of three months in a previous study, showing reduced levels of circulating plasmablasts, and naïve and transitional B cells^17^. Although the exact mechanisms remain to be unraveled, very early-life microbiota-host crosstalk at the intestinal mucosa appears to imprint on systemic immunity, including vaccine responses.

Mode of delivery and breastfeeding are important drivers of early-life *Bifidobacterium* and *E. coli* abundance^13,24,41^, whereas antibiotic treatment in the neonatal period has shown to dramatically reduce these bacteria^42^. Our results suggest that early-life microbiota mediate the relationship between mode of delivery and anti-pneumococcal vaccine responses, further emphasizing the importance of discouraging the increasing application of C-section in the absence of medical urgency to preserve the microbiota-immune axis in infants. Similarly, antibiotic-induced microbiota disruption may lead to reduced vaccine responses^7,21^. Preterm infants have also been shown to generate lower antibody levels following vaccination compared to term-born controls^43^. In our healthy, term-born cohort, very few infants required antibiotic treatment in the first weeks of life, and further studies are required to compare our findings to (preterm) infants who received antibiotics as neonates.

We observed stronger associations of specific gut colonization patterns in early life with antibody responses to pneumococcal vaccination than with antibody responses to meningococcal vaccination. Furthermore, antibody responses against pneumococcal serotypes were not correlated to those against MenC, suggesting that early-life microbe-mediated immune modulation might be antigen-specific. A more likely explanation is that pneumococcal and meningococcal vaccinations are administered at different ages. When meningococcal vaccination is administered at 14 months of age, the immune system has been exposed to other factors, and is already more mature and possibly more resilient to microbiota-related cues than when the first pneumococcal vaccination is administered at two months of age^16^. Notably, we associated higher abundances of members of the Lachnospiraceae family, including butyrate-producing taxa, with higher anti-meningococcal antibody responses. The abundance of these bacteria in the gut typically increases following the cessation of breastfeeding^41,44^, and are generally found to be beneficial for the developing immune system^45^.

Our findings lay a foundation for studies investigating interventions that modulate the infant gut microbiota to improve vaccine immunogenicity. Perturbed gut microbial colonization patterns likely contribute to reduced vaccine effectiveness across certain populations and settings^9^. Methods to modulate the gut microbiota following perturbations such as C-section birth are being investigated, and range from probiotic administration^46^ to maternal fecal microbiota transplants^47^, but it remains unknown if such interventions confer any long-term health benefits including enhanced vaccine immunogenicity. Our findings also suggest that different interventions may be indicated for vaccinations given earlier in life compared to later in life, and this should be considered in future studies.

Strengths of our work include the dense sampling at different timepoints, especially in the beginning of life. The extensively documented epidemiological data and microbiota composition of our cohort allowed us to establish associations between gut microbiota and vaccine responses in healthy infants. Furthermore, with the sensitive MIA technology, we could accurately measure antigen-specific antibody concentrations, even in very low volumes of saliva. Limitations of our work include using saliva for antibody measurements rather than serum. However, IgG concentrations in saliva were shown to correlate with serum concentrations^48^, and are, therefore, a valid proxy for systemic IgG. Furthermore, while pneumococcal and meningococcal vaccination protect from infection primarily through neutralizing IgG, we did not assess other parameters of immunity such as IgA, antibody affinity, and T cell responses. Future studies could employ a systems biology approach to obtain a complete overview of the mechanisms that underlie interindividual variation in vaccine responses^2,49^. Our observational study was also not primarily designed to study relationships between drivers, microbes and health outcomes such as antibody responses to vaccination, which limited our power to detect significant associations. Finally, the time between vaccination of the infants and antibody measurement was variable, which we corrected for in our analyses, but may still have affected our results.

In conclusion, we demonstrate that mode of delivery-associated differences in the gut microbiota in the first weeks of life, including differences in *E. coli* and *Bifidobacterium* abundances, are associated with anti-pneumococcal and anti-MenC IgG responses to vaccination. Incorporating antibody responses to vaccination as a parameter in future trials of early-life microbiota modulation could offer opportunities to assess beneficial outcomes on the microbe-mediated training of the immune system. Improved understanding of the microbial factors driving immune maturation and vaccine immunogenicity is key to improve vaccine performance and combat infectious diseases in children.

## Data Availability

Sequence data that support the findings of this study have been deposited in the NCBI Sequence Read Archive (SRA) database with BioProject ID PRJNA481243, and PRJNA555020.

https://www.ncbi.nlm.nih.gov/bioproject/PRJNA481243/

https://www.ncbi.nlm.nih.gov/bioproject/?term=PRJNA555020

## Acknowledgements

The authors are indebted to all the participating children and their families. We thank all the members of the research team of the Spaarne Gasthuis Academy, the laboratory staff, and the Streeklaboratorium Haarlem. We are grateful to Belinda van ‘t Land from Nutricia for providing some of the reagents.

## Author contributions

D.B., M.A. van H., and E.A.M.S conceived and designed the study. M.A. van H. was involved in enrolling the participants. M.L.J.N.C., F. de H., and G.A.M.B. were responsible for the execution and quality control of the laboratory work. E.M. de K., M.R., D.B., and S.F. analyzed the data. E.M. de K.,

D. van B., M.A. van H., E.A.M.S., D.B., and S.F. wrote the paper. All authors significantly contributed to interpreting the results, critically revised the manuscript for important intellectual content, and approved the final manuscript. E.M. de K., M.R., and M.L.J.N.C. have verified the microbiome and participant data. E.M. de K., and F. de H. have verified the antibody data.

## Data availability

Sequence data that support the findings of this study have been deposited in the NCBI Sequence Read Archive (SRA) database with BioProject ID PRJNA481243, and PRJNA555020. Deidentified participant data including antibody measurements, and data dictionaries can be made available after approval of a proposal.

