## appendix for "Microbiota development in early life mediates association between mode of delivery and vaccine responses"

**Supplementary Appendix**

### Extended Methods

#### Sample collection

Saliva for antibody measurement was collected during home visits at the ages of 12 and 18 months. An absorbent sponge (Malvern Medical Developments Ltd., Worcester, UK) was rubbed on the gums, cheek pouches and tongue for 1 minute. Saliva was immediately transferred to a tube containing EDTA (BD Vacutainer, New Jersey, USA) with protease inhibitor (Roche, Basel, Switzerland). Fecal samples for gut microbiota profiling were collected by the parents prior to each visit using sterile containers, and were directly stored in the home freezer, until collection by research personnel. Saliva and feces were transported on dry ice and stored at -80˚C awaiting subsequent analyses.

#### IgG measurements

Antigen-specific IgG against the capsular polysaccharides of pneumococcal vaccine serotypes 1, 4, 5, 6B, 7F, 9V, 14, 18C, 19F and 23F was measured in saliva obtained at 12 months, and IgG against MenC polysaccharide in saliva obtained at 18 months. Antibodies were quantified using fluorescent bead-based multiplex immunoassays (MIA) as previously described[1–3]. Carboxylated microspheres (Luminex, Austin, TX) were coated with the respective polysaccharide antigens. To this end, antigens were first linked to Poly-L-lysine, and then the complex was bound to the microspheres in a reaction using EDC with sulpho-NHS. Standard reference sera with previously assigned concentrations of serotype-specific IgG were an in-house intravenous immunoglobulin (IVIG) for pneumococcal serotypes (Sanquin, Amsterdam, The Netherlands), calibrated on the WHO international standard 007sp (NIBSC), and CDC1992 for MenC (NIBSC, Ridge, United Kingdom)[4]. Saliva was thawed and centrifuged, and supernatants were diluted 1:2 and 1:10 using phosphate buffered saline (PBS; pH=7·2) with 5% antibody-depleted human serum (Valley Biomedical, Winchester, VA) and with 15 ug/ml multi cell wall polysaccharide (Statens Serum Insititut, Copenhagen, Denmark). From each dilution, 25 µl was mixed with an equal volume of beads. R-phycoerythrin conjugated goat anti-human IgG solution diluted 1:200 (Jackson ImmunoResearch, West Grove, PA) was added to each well. Analysis of the beads was performed on a BioPlex 200 apparatus using the BioPlex software package version 6·2 (Bio-Rad Laboratories, Hercules, CA). IgG concentrations were determined based on averaging results of both dilutions. When the concentrations differed more than twofold (coefficient of variation >47%), the result of the 1:10 dilution was used when in standard range. IgG concentrations were expressed in ng/ml. IgG concentrations below the lower limit of detection, which ranged from 0·08 ng/ml for pneumococcal serotype 4 to 0·37 ng/ml for pneumococcal serotype 14, and was 0·21 ng/ml for MenC, were set at half the lower limit of detection.

#### Dirichlet multinomial mixture models

Dirichlet multinomial mixture models were used to group infants into community state types (CSTs) based on their gut microbiota composition (*DirichletMultinomial*[5]) (limited to OTUs present in at least 10% of samples). The optimal number of CSTs was set at the number of Dirichlet components representing optimal model fit, testing a range of 1 to 7 components. Model fit was based on the Laplace approximation to the negative log model, where a lower value indicates a better fit. Differences in the gut microbial community composition according to CST were evaluated using permutational analysis of variance (PERMANOVA) (*vegan*[6]).

### Supplementary Figures

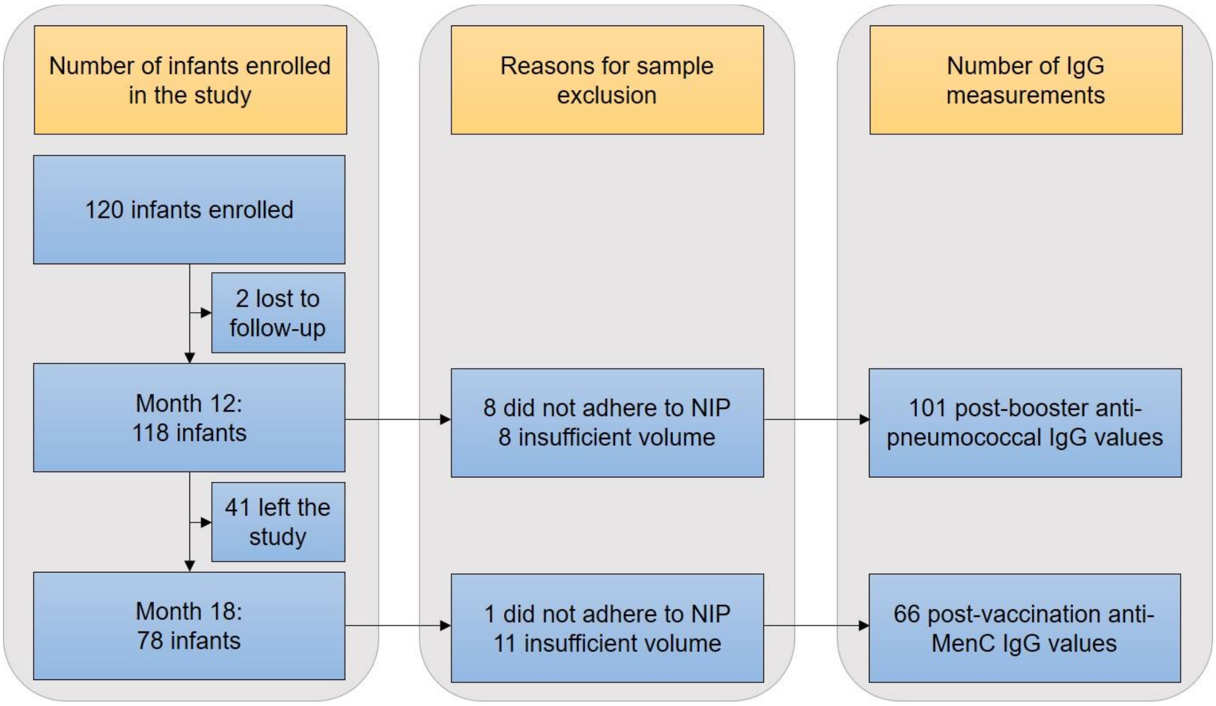

**Figure S1. Flowchart**
Saliva was collected from healthy infants at the ages 12 months and 18 months. Anti-pneumococcal IgG was measured at 12 months, and anti-meningococcal IgG at 18 months. IgG measurements were excluded from the analysis if infants did not receive their vaccinations in time, or if the saliva sample did not have a sufficient volume for laboratory analysis. *Abbreviations:* NIP = national immunization program.

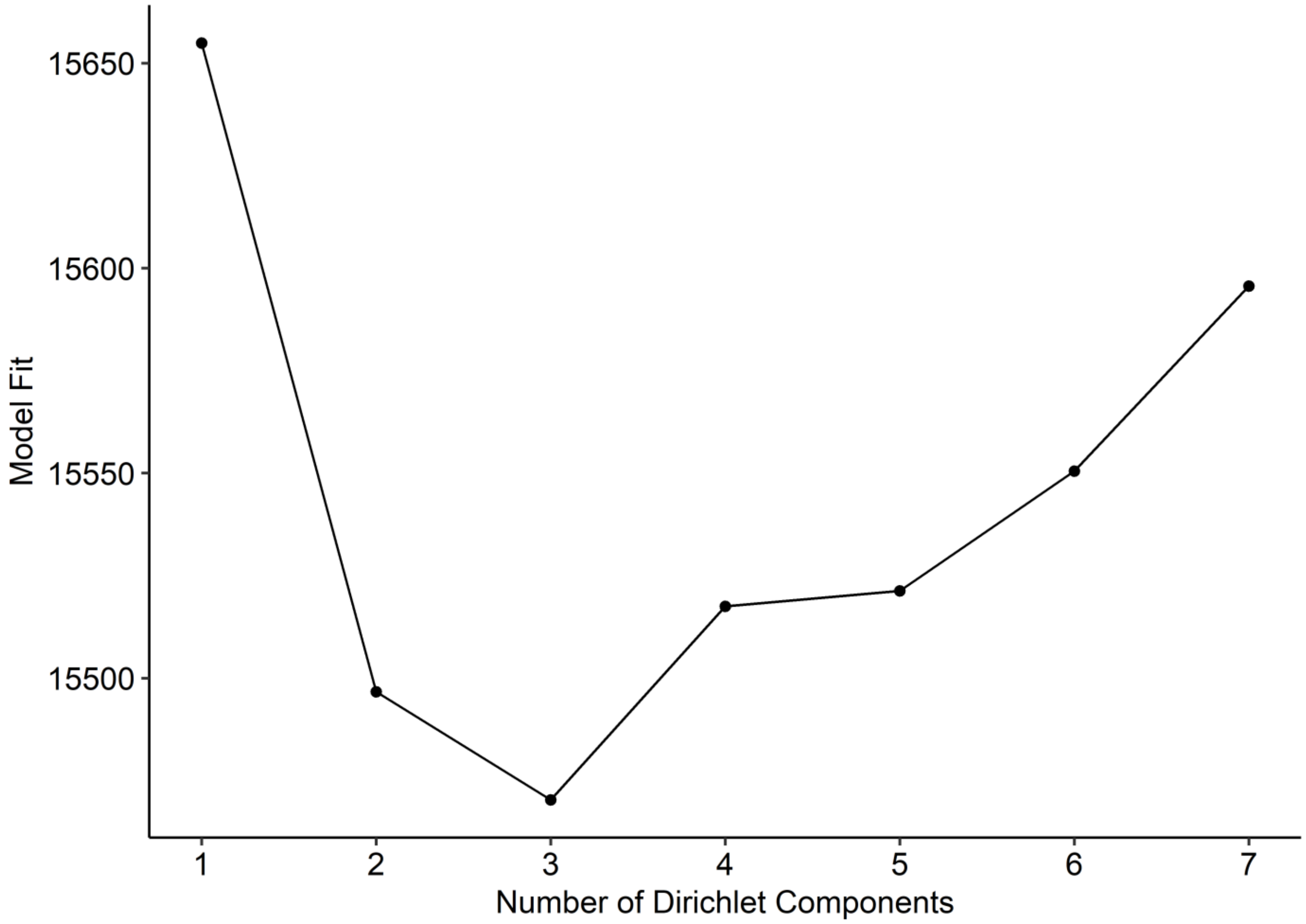

**Figure S2. Dirichlet multinomial mixture model fit.**
Dirichlet multinomial mixture model identified 3 compositionally distinct community state types (CST) as the best model fit at the week 1 timepoint. Model fit was based on the Laplace approximation to the negative log model where a lower value indicates a better model fit.

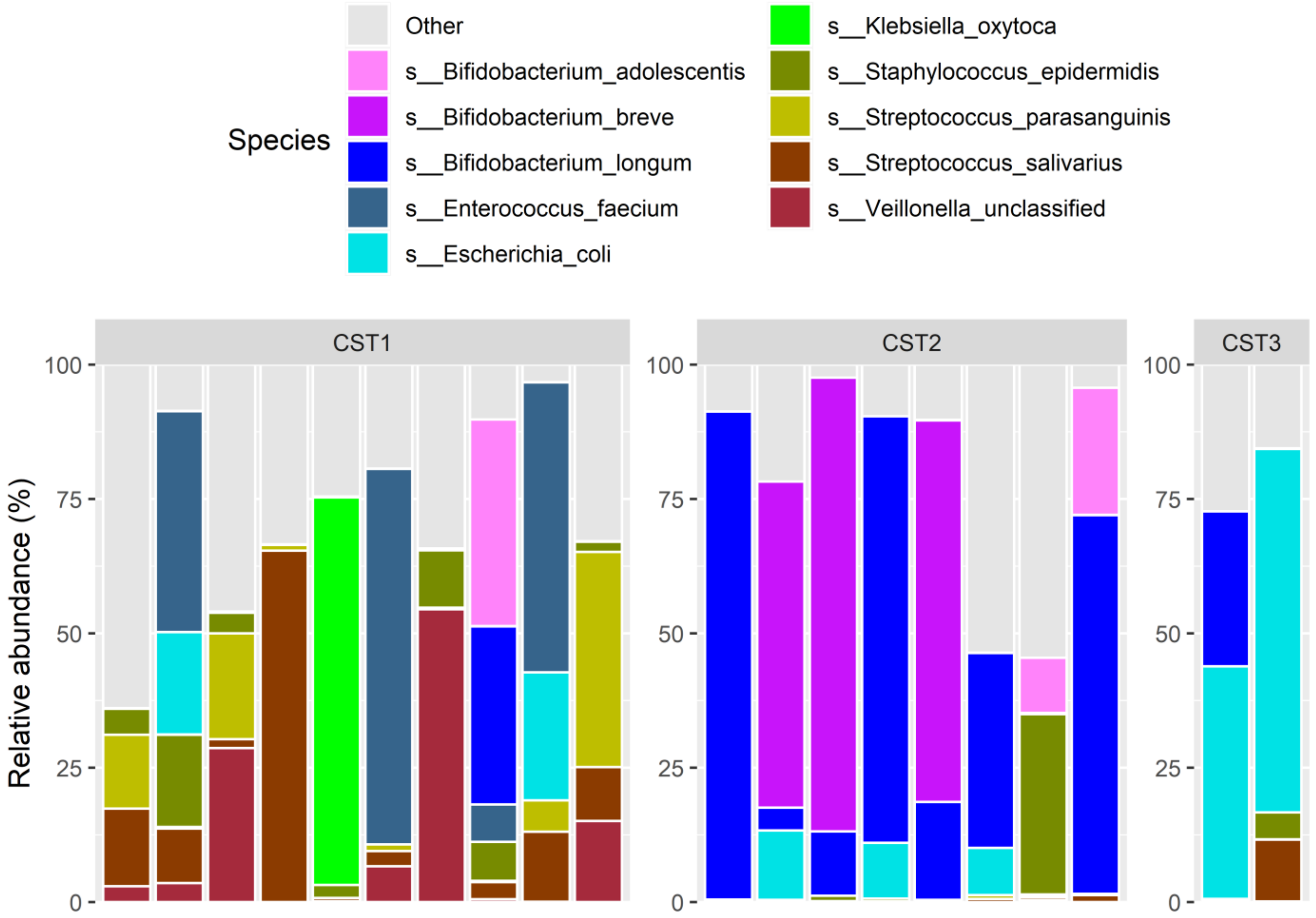

**Figure S3. Species-level composition of community state types at week 1 of age.**
Relative abundances of the top 10 species in 20 week 1 samples, determined by whole genome sequencing. Samples were ordered by week 1 community state type (CST).

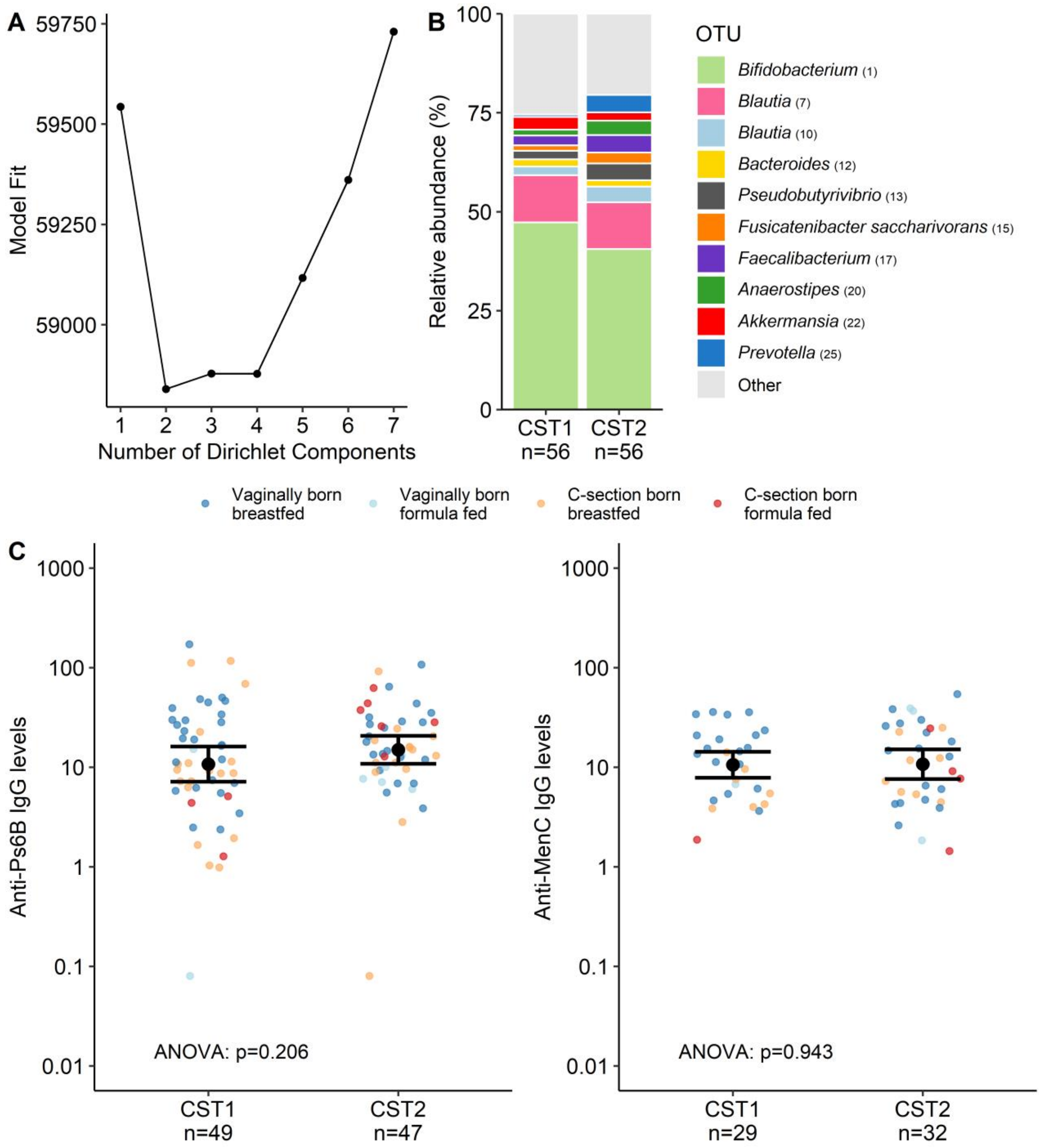

**Figure S4. Community state types at 12 months of age.**
(A) Dirichlet multinomial mixture model identified 2 compositionally distinct community state types (CST) as the best model fit at the month 12 timepoint. Model fit was based on the Laplace approximation to the negative log model where a lower value indicates a better model fit. (B) Relative abundances of the top 10 OTUs per community state type (CST) defined at 12 months of age. (C) Month 12 CSTs are plotted against anti-Ps6B IgG concentrations as representative serotype (left) and anti-MenC IgG concentrations (right). Dots are colored according to mode of delivery and feeding type from birth. Black dots and error bars represent geometric mean concentrations with 95% confidence intervals. Significance was assessed using ANOVA on log-transformed IgG concentrations followed by post-hoc Tukey tests adjusting for multiple comparisons using the Benjamini-Hochberg procedure, also correcting for time between vaccination and IgG measurements. *Abbreviations*: OTU = operational taxonomic unit.

### Supplementary Tables

**Table S1. Validation of associations between early-life characteristics and anti-pneumococcal IgG concentrations following vaccination.**

| | Formula feeding vs breastfeeding | | Vaginal birth vs C-section birth | | Female vs male | | AB vs no AB in first 3 months | | Pets vs no pets in household | | Formula feeding * vaginal birth | |
|---|---|---|---|---|---|---|---|---|---|---|---|---|
| Ps | β (95% CI) | p-value | β (95% CI) | p-value | β (95% CI) | p-value | β (95% CI) | p-value | β (95% CI) | p-value | β (95% CI) | p-value |
| 1 | 0.57 (-0.19-1.33) | 0.142 | 0.48 (0.01-0.96) | 0.046 | -0.09 (-0.52-0.34) | 0.682 | -0.26 (-0.91-0.39) | 0.429 | -0.19 (-0.62-0.24) | 0.384 | -1.87 (-3.00- -0.73) | 0.002 |
| 4 | -0.17 (-0.82-0.47) | 0.596 | 0.26 (-0.14-0.66) | 0.194 | -0.01 (-0.37-0.36) | 0.971 | -0.29 (-0.83-0.26) | 0.302 | -0.38 (-0.74- -0.02) | 0.041 | -0.80 (-1.76-0.16) | 0.100 |
| 5 | 0.35 (-0.27-0.97) | 0.263 | 0.49 (0.11-0.87) | 0.013 | 0.06 (-0.29-0.40) | 0.751 | -0.25 (-0.78-0.27) | 0.340 | -0.15 (-0.50-0.20) | 0.394 | -1.37 (-2.28- -0.45) | 0.004 |
| 7F | 0.35 (-0.29-1.00) | 0.281 | 0.41 (0.01-0.81) | 0.045 | 0.38 (0.02-0.75) | 0.040 | -0.32 (-0.87-0.23) | 0.250 | -0.25 (-0.62-0.11) | 0.171 | -1.46 (-2.42- -0.50) | 0.003 |
| 9V | 0.36 (-0.26-0.99) | 0.246 | 0.36 (-0.02-0.75) | 0.065 | 0.03 (-0.32-0.38) | 0.874 | -0.40 (-0.92-0.13) | 0.140 | -0.19 (-0.54-0.16) | 0.291 | -1.23 (-2.16- -0.31) | 0.009 |
| 14 | 0.41 (-0.31-1.12) | 0.262 | 0.32 (-0.12-0.76) | 0.154 | 0.16 (-0.24-0.57) | 0.422 | -0.23 (-0.84-0.37) | 0.444 | -0.37 (-0.77-0.03) | 0.070 | -1.50 (-2.56- -0.44) | 0.006 |
| 18C | -0.49 (-1.23-0.25) | 0.191 | 0.05 (-0.40-0.51) | 0.820 | 0.09 (-0.33-0.50) | 0.674 | -0.72 (-1.34--0.09) | 0.025 | -0.52 (-0.94--0.11) | 0.014 | -0.28 (-1.38-0.81) | 0.610 |
| 19F | 0.27 (-0.44-0.99) | 0.452 | 0.29 (-0.16-0.73) | 0.203 | 0.16 (-0.24-0.56) | 0.434 | -0.53 (-1.14-0.08) | 0.087 | -0.29 (-0.69-0.11) | 0.154 | -1.32 (-2.38- -0.26) | 0.015 |
| 23F | 0.10 (-0.60-0.80) | 0.778 | 0.39 (-0.04-0.83) | 0.076 | -0.01 (-0.40-0.39) | 0.978 | -0.40 (-0.99-0.20) | 0.187 | -0.26 (-0.66-0.13) | 0.186 | -0.79 (-1.83-0.25) | 0.135 |

*All analyses were corrected for time between vaccination and IgG measurement.*
*Abbreviations: Ps = pneumococcal serotype; C-section = caesarean section; AB = antibiotics.*

**Table S2. Validation of associations between Bray-Curtis similarity and anti-pneumococcal IgG levels.**

| Ps | Time interval | Coefficient | Adjusted p-value |
|---|---|---|---|
| 1 | d1-w1 | 1.20 | 0.065 |
| | w1-w2 | 0.86 | 0.065 |
| 4 | d1-w1 | 1.07 | 0.055 |
| | w1-w2 | 0.75 | 0.055 |
| 5 | d1-w1 | 1.00 | 0.040 |
| | w1-w2 | 0.80 | 0.040 |
| 7F | d1-w1 | 0.72 | 0.162 |
| | w1-w2 | 0.96 | 0.024 |
| 9V | d1-w1 | 1.22 | 0.011 |
| | w1-w2 | 1.02 | 0.009 |
| 14 | d1-w1 | 1.25 | 0.023 |
| | w1-w2 | 1.25 | 0.006 |
| 18C | d1-w1 | 1.77 | 0.002 |
| | w1-w2 | 0.77 | 0.083 |
| 19F | d1-w1 | 1.14 | 0.056 |
| | w1-w2 | 0.41 | 0.307 |
| 23F | d1-w1 | 1.01 | 0.077 |
| | w1-w2 | 1.15 | 0.023 |

*All analyses were corrected for time between vaccination and IgG measurement.*
*Abbreviations: Ps = pneumococcal serotype; d1=day 1; w1=week 1; w2=week 2.*

**Table S3. Validation of association between week 1 CST and IgG concentrations against pneumococcal serotypes.**

| | ANOVA | Post-hoc Tukey tests | | | | | |
|---|---|---|---|---|---|---|---|
| | All CSTs | CST2 vs CST1 | | CST3 vs CST1 | | CST3 vs CST2 | |
| Ps | p-value | estimate | adjusted p-value | estimate | adjusted p-value | estimate | adjusted p-value |
| 1 | 0.043 | 0.530 | 0.126 | 0.748 | 0.105 | 0.218 | 0.829 |
| 4 | 0.062 | 0.421 | 0.160 | 0.572 | 0.157 | 0.151 | 0.881 |
| 5 | 0.031 | 0.507 | 0.047 | 0.438 | 0.279 | -0.069 | 0.970 |
| 7F | 0.084 | 0.460 | 0.099 | 0.366 | 0.443 | -0.095 | 0.949 |
| 9V | 0.049 | 0.433 | 0.104 | 0.488 | 0.204 | 0.055 | 0.981 |
| 14 | 0.056 | 0.513 | 0.104 | 0.563 | 0.221 | 0.050 | 0.988 |
| 18C | 0.050 | 0.536 | 0.098 | 0.562 | 0.242 | 0.026 | 0.997 |
| 19F | 0.386 | 0.270 | 0.483 | 0.283 | 0.645 | 0.013 | 0.999 |
| 23F | 0.056 | 0.614 | 0.058 | 0.330 | 0.627 | -0.284 | 0.720 |

*All analyses were corrected for time between vaccination and IgG measurement.*
*Abbreviations: CST = community state type; Ps = pneumococcal serotype.*

**Table S4. CST as a mediator between mode of birth and anti-Ps6B and anti-MenC IgG concentrations.**

| | Anti-Ps6B IgG | | | | Anti-MenC IgG | | | |
|---|---|---|---|---|---|---|---|---|
| | Model without CST | | Model with CST | | Model without CST | | Model with CST | |
| | Coefficient (95% CI) | p-value | Coefficient (95% CI) | p-value | Coefficient (95% CI) | p-value | Coefficient (95% CI) | p-value |
| (Intercept) | 2.13 (1.71-2.55) | <0.0001 | 2.05 (1.62-2.48) | <0.0001 | 2.03 (1.71-2.35) | <0.0001 | 1.93 (1.59-2.27) | <0.0001 |
| Vaginal birth | 0.53 (-0.02-1.08) | 0.060 | 0.04 (-0.74-0.83) | 0.911 | 0.52 (0.12-0.91) | 0.012 | 0.68 (0.14-1.22) | 0.015 |
| CST2 vs 1 | *NA* | *NA* | 0.72 (-0.12-1.57) | 0.091 | *NA* | *NA* | -0.14 (-0.69-0.40) | 0.602 |
| CST3 vs 1 | *NA* | *NA* | 0.60 (-0.25-1.45) | 0.165 | *NA* | *NA* | 0.48 (-0.11-1.06) | 0.107 |

*All analyses were corrected for time between vaccination and IgG measurement.*
*Abbreviations: CST = community state type; NA = not applicable.*

**Table S5. Differentially abundant OTUs in the first 2 months of life between infants with high vs. low anti-Ps6B IgG levels**

| OTU | Interval number | Interval start | Interval end | Area | Association | p-value | Adjusted p-value | Validated for Ps*: |
|---|---|---|---|---|---|---|---|---|
| *Bifidobacterium (1)* | interval:1 | 0 | 5 | 7.2 | Above median | 0.013 | 0.027 | 1, 4, 5, 7F, 9V, 19F, 23F |
| *Escherichia coli (2)* | interval:1 | 0 | 41 | 82.0 | Above median | 0.003 | 0.013 | 1, 4, 5, 7F, 9V, 18C, 23F |
| *Ruminococcus gnavus (9)* | interval:1 | 0 | 16 | 20.5 | Above median | 0.016 | 0.031 | 7F |
| *Pseudobutyrivibrio (13)* | interval:1 | 0 | 20 | -14.3 | Below median | 0.050 | 0.071 | 1, 4, 5, 7F, 9V, 23F |
| *Anaerostipes (20)* | interval:1 | 0 | 17 | -8.6 | Below median | 0.064 | 0.084 | 5, 7F, 9V |
| *Clostridium sensu stricto 1 (21)* | interval:1 | 0 | 31 | -46.0 | Below median | 0.023 | 0.040 | 5, 7F, 23F |
| *Prevotella (25)* | interval:1 | 0 | 23 | -22.4 | Below median | 0.001 | 0.010 | 1, 4, 5, 7F, 9V, 14, 18C, 19F, 23F |
| *Streptococcus pyogenes (26)* | interval:1 | 0 | 33 | -21.4 | Below median | 0.013 | 0.027 | 1, 9V, 14 |
| *Dorea (32)* | interval:1 | 0 | 4 | -5.2 | Below median | 0.070 | 0.089 | 4, 5, 7F, 9V |
| *Bacteroides (53)* | interval:1 | 0 | 48 | 42.7 | Above median | 0.002 | 0.010 | 1, 4, 5, 7F, 9V, 19F, 23F |
| *Streptococcus (55)* | interval:1 | 0 | 43 | -50.3 | Below median | 0.024 | 0.040 | 1, 4, 9V |
| *Lactococcus lactis (80)* | interval:1 | 0 | 33 | -35.7 | Below median | 0.001 | 0.010 | 1, 5, 7F, 9V, 14, 18C, 19F, 23F |
| *Bifidobacterium (147)* | interval:1 | 0 | 19 | 18.0 | Above median | 0.002 | 0.010 | 1, 4, 5, 7F, 19F, 23F |
| *Escherichia/Shigella (185)* | interval:1 | 0 | 36 | 46.7 | Above median | 0.002 | 0.010 | 1, 7F, 18C |
| *Pseudomonas fluorescens (236)* | interval:1 | 0 | 5 | -3.7 | Below median | 0.088 | 0.099 | 5, 7F, 9V |
| *Bacillales (255)* | interval:1 | 0 | 39 | -38.7 | Below median | 0.001 | 0.010 | 4, 5, 7F, 9V, 19F, 23F |
| *Enterococcus (256)* | interval:1 | 0 | 45 | -48.8 | Below median | 0.002 | 0.010 | 1, 4, 5, 7F, 9V, 18C, 19F, 23F |
| *Enterobacteriaceae (345)* | interval:1 | 0 | 15 | 7.4 | Above median | 0.020 | 0.038 | |
| *Staphylococcaceae (382)* | interval:1 | 0 | 58 | -52.1 | Below median | 0.001 | 0.010 | 4, 5, 7F, 9V, 19F, 23F |
| *Streptococcus gallolyticus (18)* | interval:1 | 2 | 44 | 45.0 | Above median | 0.013 | 0.027 | 14 |
| *Bacteroides (433)* | interval:1 | 3 | 8 | 2.0 | Above median | 0.023 | 0.040 | 4, 9V, 19F, 23F |
| *Streptococcus (502)* | interval:1 | 3 | 49 | -42.3 | Below median | 0.002 | 0.010 | 1, 5, 7F, 9V, 23F |
| *Bacteroides (19)* | interval:1 | 4 | 47 | 58.0 | Above median | 0.021 | 0.039 | 1, 4, 5, 7F, 9V, 18C, 19F, 23F |

| | | | | | | | | |
|---|---|---|---|---|---|---|---|---|
| *Streptococcus (189)* | interval:1 | 4 | 26 | -26.4 | Below median | 0.007 | 0.019 | 1, 4, 5, 9V |
| *Gardnerella (333)* | interval:1 | 11 | 60 | -51.8 | Below median | 0.004 | 0.014 | 1, 5, 9V, 14, 18C |
| *Veillonella (366)* | interval:1 | 11 | 62 | -44.3 | Below median | 0.004 | 0.014 | 1, 5, 7F, 9V, 14, 23F |
| *Bacteroides (35)* | interval:1 | 12 | 47 | 42.3 | Above median | 0.009 | 0.022 | 1, 4, 5, 18C, 19F, 23F |
| *Bilophila wadsworthia (136)* | interval:1 | 14 | 70 | 52.7 | Above median | 0.006 | 0.018 | 1, 4, 5, 9V, 14, 23F |
| *Subdoligranulum (38)* | interval:2 | 15 | 30 | 13.4 | Above median | 0.006 | 0.018 | 1, 4, 14 |
| *Bifidobacterium bifidum (11)* | interval:1 | 16 | 70 | -101.4 | Below median | 0.026 | 0.041 | 9V, 14, 18C, 19F |
| *Rothia (113)* | interval:1 | 17 | 29 | -9.1 | Below median | 0.062 | 0.084 | 4, 9V, 23F |
| *Peptostreptococcus (168)* | interval:1 | 17 | 70 | 50.8 | Above median | 0.081 | 0.095 | 9V, 23F |
| *Bacteroides (249)* | interval:1 | 17 | 70 | 39.4 | Above median | 0.002 | 0.010 | 4, 5, 7F, 9V, 19F, 23F |
| *Streptococcus (69)* | interval:1 | 18 | 55 | -35.0 | Below median | 0.074 | 0.090 | |
| *Lactobacillus fermentum (75)* | interval:1 | 18 | 30 | -6.0 | Below median | 0.074 | 0.090 | 4 |
| *Veillonella (179)* | interval:1 | 19 | 32 | -7.3 | Below median | 0.029 | 0.045 | 1, 4, 5, 7F, 9V, 19F |
| *Enterococcus faecium (5)* | interval:1 | 20 | 23 | -3.8 | Below median | 0.046 | 0.067 | 5, 7F, 23F |
| *Bifidobacteriaceae (309)* | interval:1 | 21 | 48 | -14.5 | Below median | 0.089 | 0.099 | 4, 7F, 9V, 18C, 23F |
| *Bifidobacterium animalis (41)* | interval:1 | 22 | 64 | -54.7 | Below median | 0.015 | 0.031 | 1, 5, 9V, 14, 18C, 19F, 23F |
| *Bacteroides (48)* | interval:1 | 24 | 41 | 14.6 | Above median | 0.065 | 0.084 | 14, 23F |
| *Bacteroides (12)* | interval:1 | 26 | 70 | 81.1 | Above median | 0.007 | 0.019 | 7F, 23F |
| *Bifidobacterium (229)* | interval:1 | 29 | 70 | -40.3 | Below median | 0.005 | 0.017 | 9V, 14, 18C, 19F |
| *Blautia (10)* | interval:1 | 33 | 59 | 140.0 | Above median | 0.003 | 0.013 | 1, 4, 19F, 23F |
| *Enterobacteriaceae (242)* | interval:1 | 33 | 70 | -42.4 | Below median | 0.002 | 0.010 | 9V, 18C, 19F, 23F |
| *Klebsiella (4)* | interval:1 | 34 | 70 | -80.7 | Below median | 0.004 | 0.014 | |
| *Carnobacteriaceae (311)* | interval:1 | 38 | 70 | 22.6 | Above median | 0.061 | 0.084 | 14, 19F |
| *Bifidobacterium animalis (175)* | interval:1 | 39 | 70 | -27.0 | Below median | 0.082 | 0.095 | 14 |
| *Citrobacter sedlakii (288)* | interval:1 | 46 | 70 | -23.1 | Below median | 0.026 | 0.041 | 9V |
| *Finegoldia (203)* | interval:1 | 50 | 70 | -20.0 | Below median | 0.034 | 0.051 | |
| *Blautia (7)* | interval:1 | 57 | 70 | -23.5 | Below median | 0.012 | 0.027 | 4, 5, 7F, 9V, 18C |
| *Corynebacterium propinquum (79)* | interval:1 | 66 | 70 | -11.6 | Below median | 0.009 | 0.022 | |

*All analyses were corrected for time between vaccination and IgG measurements*

** Validation for other pneumococcal vaccine serotypes 1, 4, 5, 7F, 9V, 14, 18C, 19F and 23F was performed*

*using the same method. OTUs were considered validated for a given serotype if they were significantly differentially abundant (adjusted p<0.100) and were associated with the same response category (above or below median).*
*Abbreviations: Ps = pneumococcal serotype*

**Table S6. Differentially abundant OTUs in the first 2 months of life between infants with high vs. low anti-MenC IgG levels**

| OTU | Interval number | Interval start | Interval end | Area | Association | p-value | Adjusted p-value |
|---|---|---|---|---|---|---|---|
| *Escherichia coli (2)* | interval:1 | 0 | 13 | 23.2 | Above median | 0.062 | 0.072 |
| *Veillonella (8)* | interval:1 | 0 | 7 | -9.2 | Below median | 0.037 | 0.055 |
| *Peptostreptococcaceae (46)* | interval:1 | 0 | 7 | 9.4 | Above median | 0.021 | 0.054 |
| *Bacteroides (65)* | interval:1 | 0 | 26 | 25.8 | Above median | 0.014 | 0.054 |
| *Streptococcus (69)* | interval:1 | 0 | 14 | -18.5 | Below median | 0.023 | 0.054 |
| *Rothia (113)* | interval:1 | 0 | 43 | -52.8 | Below median | 0.027 | 0.054 |
| *Bifidobacteriaceae (299)* | interval:1 | 0 | 16 | -7.9 | Below median | 0.005 | 0.036 |
| *Lachnospiraceae (96)* | interval:1 | 1 | 51 | 47.4 | Above median | 0.003 | 0.029 |
| *Collinsella (16)* | interval:1 | 3 | 20 | 23.7 | Above median | 0.037 | 0.055 |
| *Veillonella (85)* | interval:1 | 3 | 16 | -20.9 | Below median | 0.002 | 0.029 |
| *Klebsiella (252)* | interval:1 | 4 | 12 | -10.8 | Below median | 0.001 | 0.029 |
| *Veillonella (368)* | interval:1 | 5 | 14 | -7.7 | Below median | 0.013 | 0.054 |
| *Bifidobacterium (218)* | interval:1 | 6 | 42 | -25.1 | Below median | 0.025 | 0.054 |
| *Clostridium sensu stricto 1 (24)* | interval:1 | 7 | 20 | -19.7 | Below median | 0.017 | 0.054 |
| *Escherichia Shigella (185)* | interval:1 | 10 | 24 | 12.3 | Above median | 0.043 | 0.059 |
| *Bifidobacteriaceae (309)* | interval:1 | 11 | 53 | 34.6 | Above median | 0.060 | 0.072 |
| *Enterococcaceae (251)* | interval:1 | 13 | 39 | -28.1 | Below median | 0.012 | 0.054 |
| *Veillonella (160)* | interval:1 | 15 | 41 | -29.0 | Below median | 0.034 | 0.055 |
| *Lactobacillus (49)* | interval:1 | 21 | 55 | -47.0 | Below median | 0.062 | 0.072 |
| *Bilophila wadsworthia (136)* | interval:1 | 21 | 69 | -54.8 | Below median | 0.022 | 0.054 |
| *bacterium NLAE zl C558 (45)* | interval:1 | 26 | 69 | 54.9 | Above median | 0.068 | 0.076 |
| *Lactobacillus fermentum (75)* | interval:1 | 33 | 69 | -49.0 | Below median | 0.037 | 0.055 |
| *Lachnospiraceae (30)* | interval:1 | 35 | 69 | -39.3 | Below median | 0.023 | 0.054 |
| *Bifidobacterium breve (261)* | interval:1 | 38 | 69 | 30.6 | Above median | 0.072 | 0.077 |
| *Streptococcus salivarius (6)* | interval:1 | 40 | 69 | -42.4 | Below median | 0.028 | 0.054 |

| | | | | | | | |
|---|---|---|---|---|---|---|---|
| *Enterobacteriaceae (281)* | interval:1 | 45 | 62 | -11.9 | Below median | 0.054 | 0.071 |
| *Clostridium butyricum (33)* | interval:1 | 62 | 69 | 26.2 | Above median | 0.038 | 0.055 |

*All analyses were corrected for time between vaccination and IgG measurements*

**Table S7. Differentially abundant OTUs of the Lachnospiraceae family in the first 12 months of life between infants with high vs. low anti-MenC IgG levels**

| OTU | Interval number | Interval start | Interval end | Area | Association | p-value | Adjusted p-value |
|---|---|---|---|---|---|---|---|
| *Lachnospira (89)* | interval:1 | 32 | 169 | 60.8 | Above median | 0.013 | 0.052 |
| *Lachnospiraceae (30)* | interval:1 | 82 | 134 | -49.9 | Below median | 0.013 | 0.052 |
| *Pseudobutyrivibrio (132)* | interval:1 | 90 | 381 | 144.2 | Above median | 0.030 | 0.066 |
| *Blautia (67)* | interval:1 | 100 | 381 | 313.6 | Above median | 0.020 | 0.053 |
| *Fusicatenibacter saccharivorans (15)* | interval:1 | 101 | 381 | 439.7 | Above median | 0.050 | 0.080 |
| *Pseudobutyrivibrio (13)* | interval:1 | 125 | 381 | 353.8 | Above median | 0.004 | 0.036 |
| *Lachnospiraceae (306)* | interval:1 | 130 | 381 | 130.4 | Above median | 0.023 | 0.054 |
| *Roseburia (77)* | interval:1 | 155 | 381 | 284.2 | Above median | 0.019 | 0.053 |
| *Dorea (54)* | interval:1 | 156 | 381 | 185.8 | Above median | 0.075 | 0.094 |
| *Blautia (28)* | interval:1 | 204 | 286 | 230.8 | Above median | 0.027 | 0.062 |
| *Lachnospira (117)* | interval:1 | 207 | 357 | 242.1 | Above median | 0.002 | 0.021 |
| *Roseburia (133)* | interval:1 | 257 | 381 | 150.6 | Above median | 0.002 | 0.021 |
| *Blautia (47)* | interval:1 | 277 | 381 | 198.6 | Above median | 0.023 | 0.054 |
| *Moryella (71)* | interval:1 | 317 | 381 | 85.6 | Above median | 0.065 | 0.092 |
| *Blautia (28)* | interval:2 | 362 | 381 | 87.5 | Above median | 0.015 | 0.053 |

*All analyses were corrected for time between vaccination and IgG measurements*
